# Revealing antibiotic cross-resistance patterns in hospitalized patients through Bayesian network modelling

**DOI:** 10.1101/2020.07.02.20144006

**Authors:** Stacey S Cherny, Daniel Nevo, Avi Baraz, Shoham Baruch, Ohad Lewin-Epstein, Gideon Y Stein, Uri Obolski

## Abstract

**Objectives:** Microbial resistance exhibits dependency patterns between different antibiotics, termed cross-resistance and collateral sensitivity. These patterns differ between experimental and clinical settings. It is unclear whether the differences result from biological reasons or from confounding, biasing results found in clinical settings. We set out to elucidate the underlying dependency patterns between resistance to different antibiotics from clinical data, while accounting for patient characteristics and previous antibiotic usage.

**Methods:** Additive Bayesian network modelling was employed to simultaneously estimate relationships between variables in a dataset of bacterial cultures derived from hospitalized patients and tested for resistance to multiple antibiotics. Data contained resistance results, patient demographics, and previous antibiotic usage, for five bacterial species: *E. coli* (n=1054), *K. pneumoniae* (n=664), *P. aeruginosa* (n=571), CoNS (n=495), and *P. mirabilis* (n=415).

**Results:** All links between resistance to the various antibiotics were positive. Multiple direct links between resistance of antibiotics from different classes were observed across bacterial species. For example, resistance to gentamicin in *E.coli* was directly linked with resistance to ciprofloxacin (OR = 8.39, 95%CI[5.58, 13.30]) and sulfamethoxazole-trimethoprim (OR = 2.95, 95%CI[1,97, 4.51]). In addition, resistance to various antibiotics was directly linked with previous antibiotic usage.

**Conclusions:** Robust relationships among resistance to antibiotics belonging to different classes, as well as resistance being linked to having taken antibiotics of a different class, exist even when taking into account multiple covariate dependencies. These relationships could help inform choices of antibiotic treatment in clinical settings.

## Introduction

The increase in antibiotic resistance frequencies of bacterial pathogens is leading to an ever-growing burden on healthcare systems, in terms of costs for newer drugs, extended hospital stays, and increased followup visits, as well as increased cost in terms of lost productivity and mortality.^1,2^ According to a new report by the United States Centers for Disease Control, annually in the USA, 2.8 million people suffer from infections that are resistant to antibiotics, resulting in 35,000 deaths per year. They reported that multidrug-resistant *P. aeruginosa*, one of the bacterial species examined in the present paper, was estimated to cause nearly 10% of the deaths attributable to antibiotic resistant bacteria and over $750M dollars in direct healthcare costs in the USA in 2017.^3^ Hence, understanding the nature of the relationships among patient covariates and the resistance to the different drugs in the current arsenal of antibiotics is crucial.

Beyond well studied, patient-related, predictors for antibiotic resistance, such as age, patients’ independence status, previous antibiotic usage, and more,^4–6^ resistance to different antibiotics is often not independent. Cross-resistance, for instance, refers to the existence of positive associations between resistance to different antibiotics across bacterial pathogens. Such dependencies are a phenomenon known for nearly as long as antibiotics have been available,^7^ with the mechanisms behind it numerous.^8^

Cross-resistance can occur due to inherent biological attributes of bacterial pathogens, when antibiotics have similar mechanisms of action and hence shared mechanisms of resistance; cross-resistance between chemically dissimilar antibiotics can also occur due to horizontal gene transfer of genetic elements coding for resistance to multiple antibiotics.^9^ Conversely, there is also evidence of cross-sensitivity, or collateral sensitivity, where negative associations between resistance to different antibiotics are observed. However, while collateral sensitivity has been demonstrated in laboratory experiments,^10–13^ when examining cultures obtained from patients in a hospital setting, it appears that host factors induce mostly positive correlations between resistance to different antibiotics.^8^ As a result of the complex relationships of resistance among different antibiotics, examining resistance to a single antibiotic or even pairs of antibiotics is insufficient. A multivariate approach which allows multiple drug resistances as dependent variables is warranted to allow uncovering the underlying structure of the observed resistance patterns, which can include conditional dependencies between the antibiotics. One such approach, that we utilize in this paper, is additive Bayesian network (ABN) modelling.^14–16^

ABN modelling is a purely data-driven approach to inferring underlying probabilistic structure of a set of variables. By essentially searching all possible directed acyclic graphs (DAGs) linking a set of variables, evidence for potential causal links can be revealed from the data without making strong prior assumptions. An approach with few prior assumptions regarding dependencies is of great utility in this area, where resistance to many different drugs and the presence of several important covariates need to be analyzed, but no strong hypotheses regarding many of the connections are available. The method has been successfully applied in veterinary studies of disease,^17^ as well as to antibiotic resistance in animals.^18–20^ A related Bayesian approach was employed in a study of meticillin-resistant *Staphylococcus aureus* transmission in humans.^21^ However, to our knowledge, the present application is the first time ABN has been used in a study of antibiotic resistance to infer cross-resistance patterns in human infections.

In this study, we employed ABN modelling to explore the interrelationships among bacterial resistance to several antibiotics, using data from bacterial cultures obtained from a single hospital in Israel. We used ABN modeling separately on each of the five bacterial species most frequent in our dataset, to examine predictors of resistance and the interrelationships among the antibiotics tested. (See Supplementary material for an illustrative example of the advantage of ABN modelling of cross-resistance compared to traditional regression approaches.)

## Materials and methods

### Ethics

The study was approved by the Helsinki Committee of Rabin Medical Center.

### Data

We obtained data pertaining to bacterial cultures drawn in Rabin Medical Center, Israel, from 2013-05-01 to 2015-12-31. The corresponding demographics, previous hospitalizations, and previous in-hospital antibiotic usage in the year prior to the infection, of patients from whom the cultures were drawn, were also available. Bacterial cultures were tested for antibiotic resistance for an array of antibiotics which had varying rates of resistance, and results of non-susceptibility and resistance were combined into a ‘resistant’ category. Bacterial infections were considered nosocomial if infections occurred >48 hours after admission. A summary of these variables is presented in Table 1. For our analyses, we selected the five bacterial species with the largest sample sizes available in the dataset: *Escherichia coli, Klebsiella pneumoniae, Pseudomonas aeruginosa, Proteus mirabilis*, and Coagulase-negative *staphylococci* (CoNS).

**Table 1:** Descriptive statistics of patients and bacterial isolates.

|  | *E. coli*<br>(n=1054) | *K. pneumoniae*<br>(n=664) | *P. aeruginosa*<br>(n=571) | *P. mirabilis*<br>(n=415) | CoNS<br>(n=495) |
| --- | --- | --- | --- | --- | --- |
| **Age: Mean (SD)** | 76.7 (14.8) | 73.5 (15.0) | 71.7 (16.1) | 74.8 (14.1) | 72.8 (18.0) |
| **Female** | 568 (53.9%) | 251 (37.8%) | 197 (34.5%) | 131 (31.6%) | 221 (44.6%) |
| **T.F.aminoglycoside** | 142 (13.5%) | 137 (20.6%) | 120 (21.0%) | 101 (24.3%) | 90 (18.2%) |
| **T.F.fluoroquinolone** | 268 (25.4%) | 214 (32.2%) | 142 (24.9%) | 97 (23.4%) | 136 (27.5%) |
| **T.F.betalactam** | 515 (48.9%) | 468 (70.5%) | 437 (76.5%) | 271 (65.3%) | 305 (61.6%) |
| **T.F.other** | 406 (38.5%) | 354 (53.3%) | 380 (66.5%) | 237 (57.1%) | 268 (54.1%) |
| **Hosp: Mean (SD)** | 2.05 (1.29) | 2.68 (1.10) | 2.91 (1.11) | 2.91 (1.29) | 2.41 (1.19) |
| **Nosocomial** | 237 (22.5%) | 244 (36.7%) | 344 (60.2%) | 198 (47.7%) | 196 (39.6%) |
| **Polymicrobial culture** | 192 (18.2%) | 225 (33.9%) | 289 (50.6%) | 213 (51.3%) | 35 (7.1%) |
| **Amikacin** |  | 93 (14.0%) |  |  |  |
| **Ampicillin** | 858 (81.4%) |  |  |  |  |
| **Ceftazidime** |  |  | 74 (13.0%) |  |  |
| **Cefuroxime** |  |  |  | 233 (56.1%) |  |
| **Chloramphenicol** |  |  |  |  | 239 (48.3%) |
| **Ciprofloxacin** | 523 (49.6%) | 389 (58.6%) |  | 241 (58.1%) |  |
| **Erythromycin** |  |  |  |  | 328 (66.3%) |
| **Fusidic acid** |  |  |  |  | 218 (44.0%) |
| **Gentamicin** | 216 (20.5%) | 279 (42.0%) | 87 (15.2%) | 219 (52.8%) | 196 (39.6%) |
| **Imipenem** |  |  | 91 (15.9%) |  |  |
| **Ofloxacin** |  |  |  |  | 291 (58.8%) |
| **Oxacillin** |  |  |  |  | 366 (73.9%) |
| **Sulf-Trim** | 534 (50.7%) |  |  | 279 (67.2%) | 253 (51.1%) |

Note: T.F. prefix denotes having taken an antibiotic from the given class; Hosp = Log of the number of days hospitalized in the prior year plus 1; Sulf-Trim = sulfamethoxazole-trimethoprim. Antibiotics listed refer to resistance to those antibiotics.

### Statistical analysis

We selected which antibiotics to include in the analysis by keeping only those with minimal missing data and which did not reduce the number of complete cases appreciably (<10% loss). We performed some variable selection to assure stable statistical models with no perfect or near-perfect separation, by not including perfectly or near perfectly correlated antibiotics and selecting only antibiotics which contained a minimum of 3% resistance in each bacterial subsample. Variables excluded from analysis are presented in Table 2, along with their tetrachoric correlations with the relevant included variables. This resulted in analysis of between three and seven antibiotics for the five bacterial species, each analyzed separately. When constructing the ABN, the following covariates were included, in addition to the antibiotic resistance variables: demographic variables (age, sex, and days hospitalized in the previous year), binary culture type variables (nosocomial and polymicrobial), and a binary variable for antibiotic use in hospital in the previous year. Antibiotics used were first grouped into classes and the three most frequent classes across the entire dataset were included, along with whether any other antibiotics were taken which did not belong to the three largest classes. The classes were aminoglycosides, fluoroquinolones, betalactams, and other antibiotics not part of these three classes. The rationale of using binary indicators for prior treatment relied on antibiotics differing in their course lengths, due to variables such as the dosage, mechanism of action, and administration route. Due to our aggregation of antibiotics into broad classes, and missing data regarding administration route, we chose to represent the treatment data using a binary indicator. Table 1 presents summary statistics for all variables used in our models.

**Table 2:** Antibiotic resistances excluded from analysis due to redundancies or missing data, along with their tetrachoric correlations with highly-correlated included variables

| Dataset | Antibiotic removed | Antibiotic it is correlated with | Tetrachoric correlation |
| --- | --- | --- | --- |
| *E.coli* | Amikacin | Ampicillin | 0.97 |
| *E.coli* | Ceftazidime | Cefuroxime | 1.00 |
| *E.coli* | Ceftriaxone | Cefuroxime | 1.00 |
| *E.coli* | Cefuroxime | Ciprofloxacin | 0.79 |
| *E.coli* | Cefuroxime | Ofloxacin | 1.00 |
| *E.coli* | Ertapenem | Cefuroxime | 0.94 |
| *K. pneumoniae* | Ceftazidime | Cefuroxime | 1.00 |
| *K. pneumoniae* | Ceftriaxone | Cefuroxime | 1.00 |
| *K. pneumoniae* | Ertapenem | Cefuroxime | 0.94 |
| *K. pneumoniae* | Cefuroxime | Ciprofloxacin | 0.82 |
| *K. pneumoniae* | Sulf-Trim | Ciprofloxacin | 0.78 |

Data were analyzed using ABN modelling,^14,15^ with version 2.1 of the R package abn^16^ on an R 3.6.1 installation.^22^ Briefly, ABN modelling is a purely data-driven, exploratory approach, originating in machine learning, that is often used for hypothesis generation for causation among a set of variables. By essentially searching all possible directed acyclic graphs (DAGs) linking a set of variables, a model for the dependency of the variables in the data can be inferred without making strong prior assumptions. This model, depicted in a DAG, shows which variables are directly connected, or linked, via arcs, and the coefficients of these arcs are directly analogous to the adjusted odds ratios obtained from multiple logistic regression analysis. While no assumptions of causal relationships are required, we restricted the model space to disallow causal paths that made no sense for our study: (1) sex could only cause other variables and not be caused by any other variable, (2) age at testing could be caused by sex but no other variable, (3) the four variables for antibiotics given in the previous year could only be caused by age and sex and no other variable, (4) hospitalization in the prior year could not be caused by presence of nosocomial infection but could be caused by any other variable, (5) nosocomial infection could not be caused by resistance to any drug but can be caused by any other variable; and (6) polymicrobial cultures as well as all drug resistances could be caused by any of the variables. In addition, no causal paths were forced to be in the model.

There are several steps involved when implementing an ABN analysis. We first used the exact search method to determine the maximum number of parents (i.e., nodes that have an arc causing another node) needed for any child (i.e., variable caused by another variable) in the model, above which there was no improvement in likelihood, by repeatedly running the exact search and setting the maximum number of parents between one and six. Never were more than four parents needed (see Supplementary Table S1). Next, we used a parametric bootstrapping approach to correct for overfitting.^23^ This was done by simulating 1000 samples of the same size as in the original data, by feeding the ABN model chosen (which arcs were present and their parameter estimates) to the JAGS software, version 4.3.0.^24^ This produces random samples generated from the parameter estimates of the chosen model. These samples were each analyzed with ABN in exactly the same way as performed initially. Next, we examined each of the 1000 ABN models and retained only those arcs present in a majority (at least 50%) of them, a common cutoff as suggested by the ABN developers.^14^ Supplementary Table S1 contains the number of arcs present in the model both before and after bootstrapping. Finally, we ran ABN on the original dataset constraining the model to only include the selected consensus arcs.This final model was used to produce 95% Bayesian credible intervals for each of the parameter estimates, which are analogous to coefficients from logistic regression models, i.e., the OR of the effect of parent (independent) on child (dependent) variable. Due to the existence of topologies that are equivalent in terms of likelihood in Bayesian networks,^20,25–30^ we present the models’ arcs and their corresponding parameters, but do not interpret their direction.

## Results

Before fitting the ABNs, we removed antibiotic variables which were nearly identical in their resistance patterns. This yielded expected results between similar antibiotics but also some extremely high associations between antibiotics of different classes. For example, we found that in all cultures where they were tested, the betalactams ceftazidime, ceftriaxone, and cefuroxime were all perfectly inter-correlated, and ertapenem and piperacillin were nearly perfectly correlated with them. However, amikacin and ampicillin were nearly perfectly correlated in the *E.coli* dataset, yet are members of different drug classes. These and additional highly correlated pairs of antibiotics can be found in Table 2. It is important to note that the results derived below are practically identical for members of the antibiotic pairs presented in Table 2.

We present the final ABNs estimated from our data, for five bacterial species, in Figures 1 through 5. Each arc in the plots represents an estimated direct link between the variables, where adjacent numbers are the parameter estimates of the OR of this link, along with their 95% credible intervals. In addition, these values are presented in equation form in Supplementary Tables 2-6, which includes the directionality of the arcs. Notably, none of the credible intervals contained the value one, i.e. no association, although this is not a required condition by the Bayesian network structure discovery algorithm.^14^ This strengthens the notion that the connections found by our models are robust.

**Figure 1:**
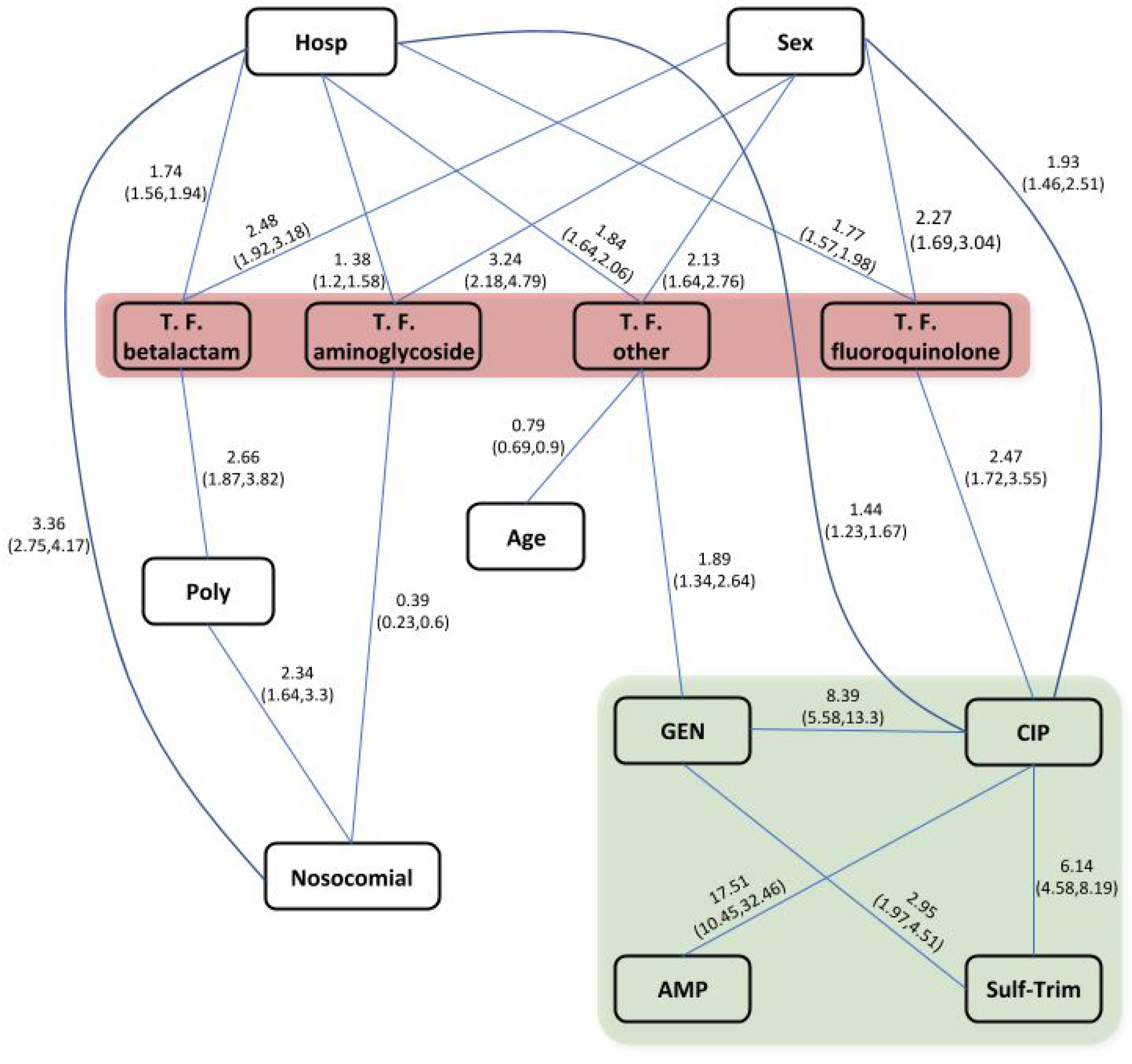
ABN model for *E. coli*. Arcs denote direct links between variables and adjacent values are ORs derived from the model, along with their 95% credible intervals (see Table S2). Nodes with antibiotic names denote resistance to those antibiotics (enclosed in the light grey rectangle [green in the color online version]). T.F. prefix denote variables indicating having taken an antibiotic from the given class in the prior year (enclosed in the dark grey rectangle [red in the color online version]); Hosp = log(days hospitalized + 1); Poly = polymicrobial culture; AMP = ampicillin; CIP = ciprofloxacin; GEN = gentamicin; Sulf-Trim = sulfamethoxazole-trimethoprim; Sex=male.

**Figure 2:**
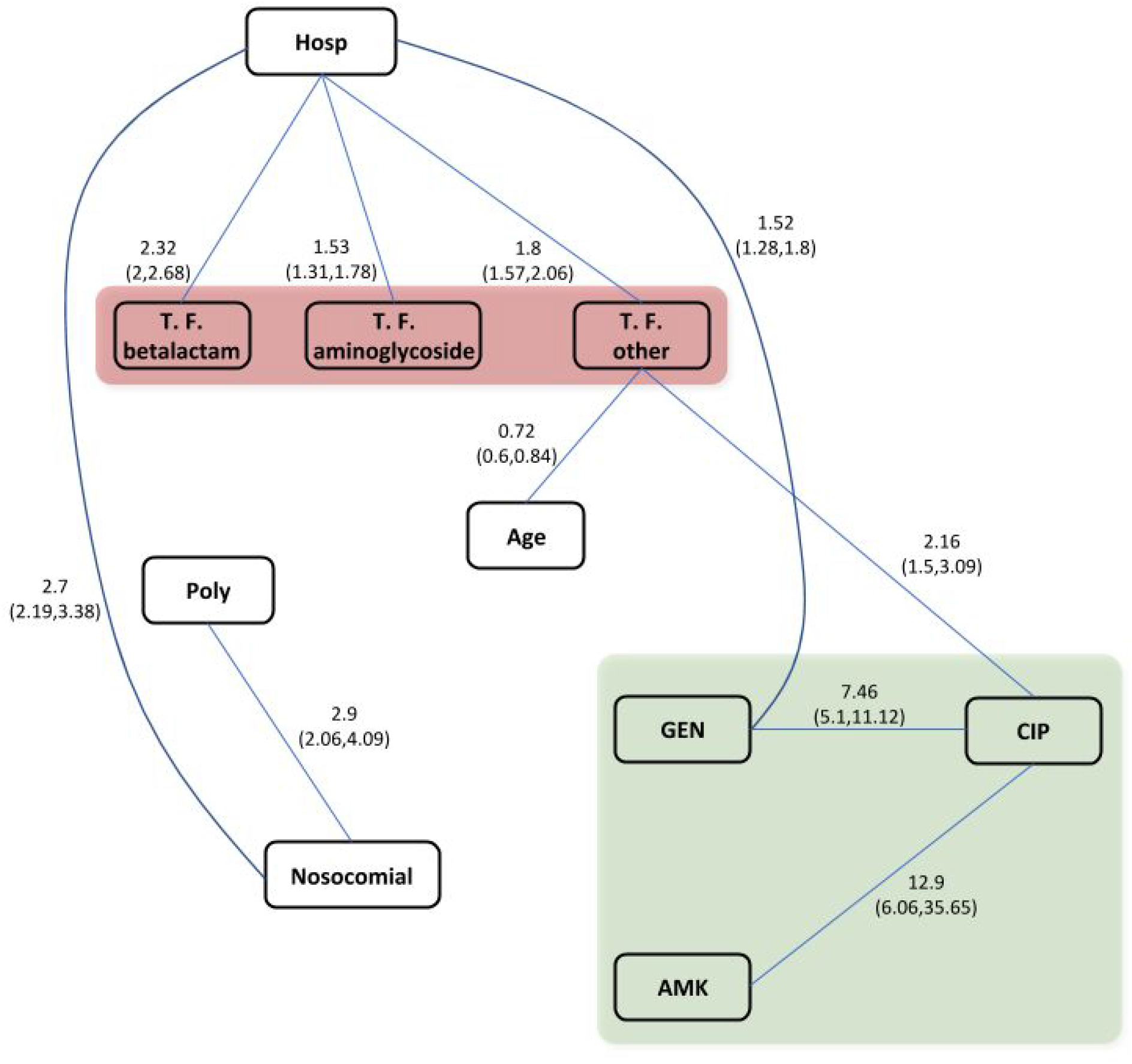
ABN model for *K. pneumoniae*. Arcs denote direct links between variables, and adjacent values are ORs derived from the model, along with their 95% credible intervals (see Table S3). Nodes with antibiotic names denote resistance to those antibiotics (enclosed in the light grey rectangle [green in the color online version]). T.F. prefix denotes variables indicating having taken an antibiotic from the given class in the prior year (enclosed in the dark grey rectangle [red in the color online version]); Hosp = log(days hospitalized + 1); Poly = polymicrobial culture; AMK = amikacin; CIP = ciprofloxacin; GEN = gentamicin.

**Figure 3:**
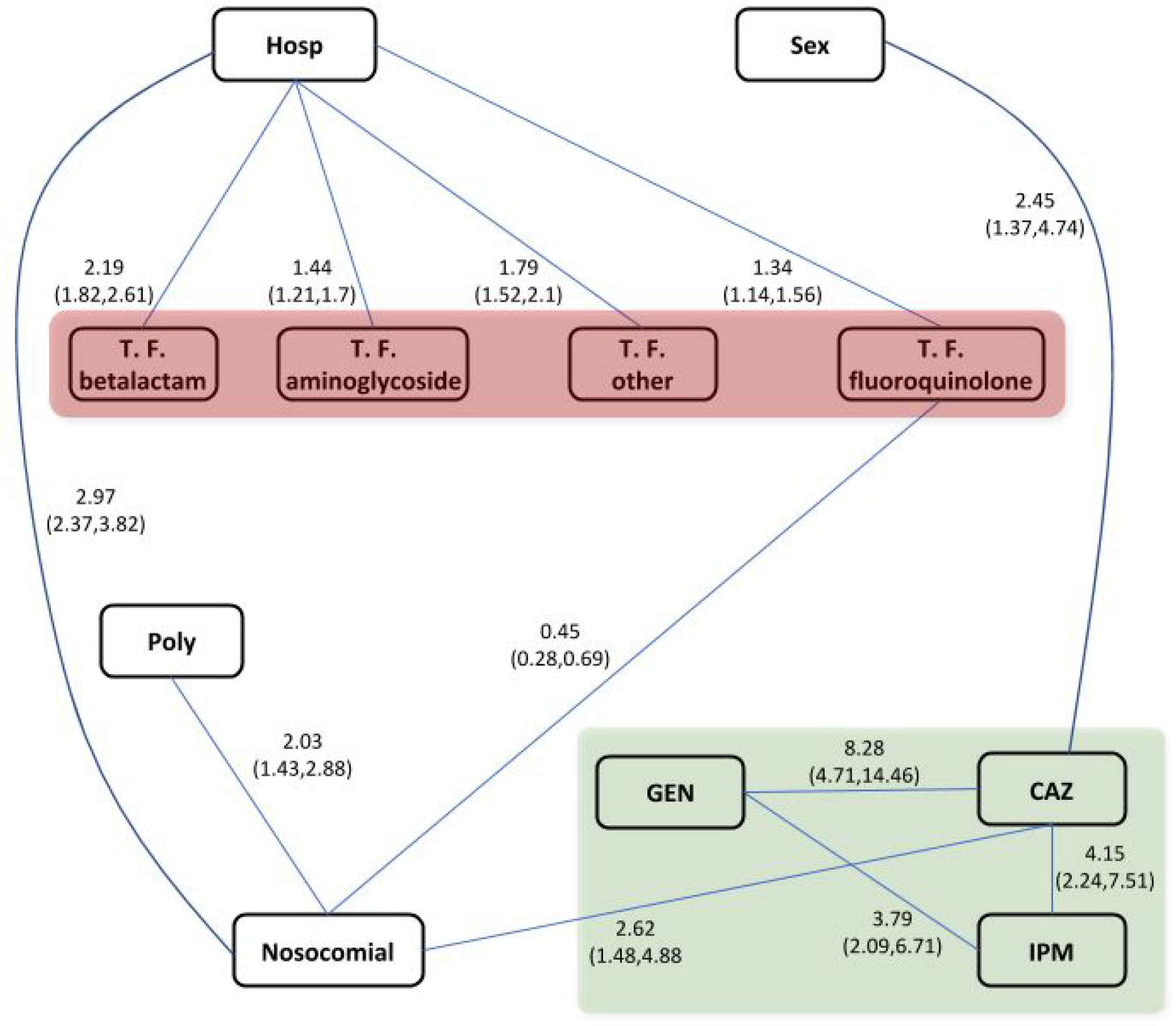
ABN model for *P. aeruginosa*. Arcs denote direct links between variables, and adjacent values are ORs derived from the model, along with their 95% credible intervals (see Table S4). Nodes with antibiotic names denote resistance to those antibiotics (enclosed in the light grey rectangle [green in the color online version]). T.F. prefix denotes variables indicating having taken an antibiotic from the given class in the prior year (enclosed in the dark grey rectangle [red in the color online version]); Hosp = log(days hospitalized + 1); Poly = polymicrobial culture; CAZ = ceftazidime; GEN = gentamicin; IPM = imipenem; Sex=male.

**Figure 4:**
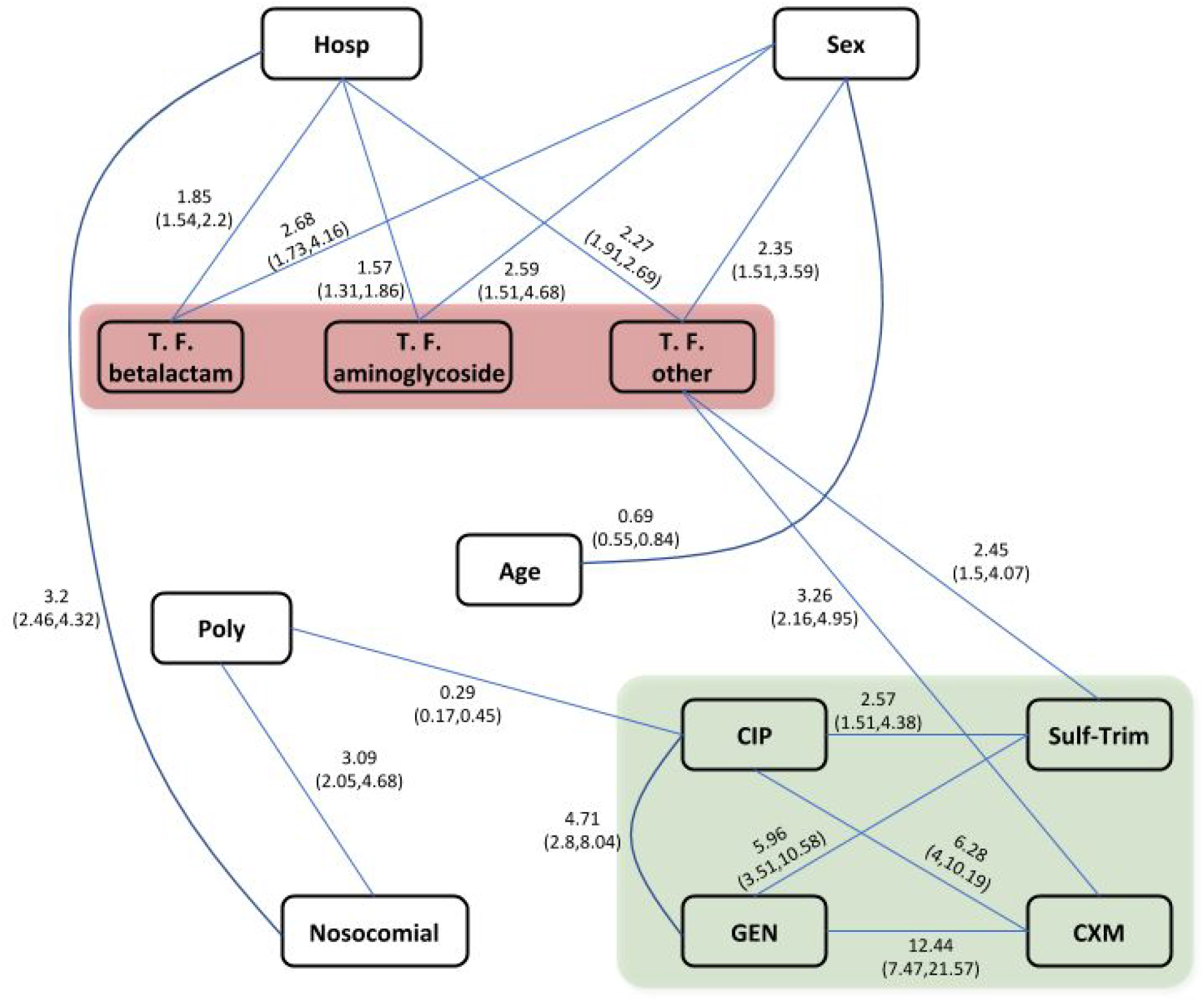
ABN model for *P. mirabilis*. Arcs denote direct links between variables, and adjacent values are ORs derived from the model, along with their 95% credible intervals (see Table S5). Nodes with antibiotic names denote resistance to those antibiotics (enclosed in the light grey rectangle [green in the color online version]). T.F. prefix denotes variables indicating having taken an antibiotic from the given class in the prior year (enclosed in the dark grey rectangle [red in the color online version]); Hosp = log(days hospitalized + 1); Poly = polymicrobial culture; CXM = cefuroxime; CIP = ciprofloxacin; GEN = gentamicin; Sulf-Trim = sulfamethoxazole-trimethoprim; Sex=male.

**Figure 5:**
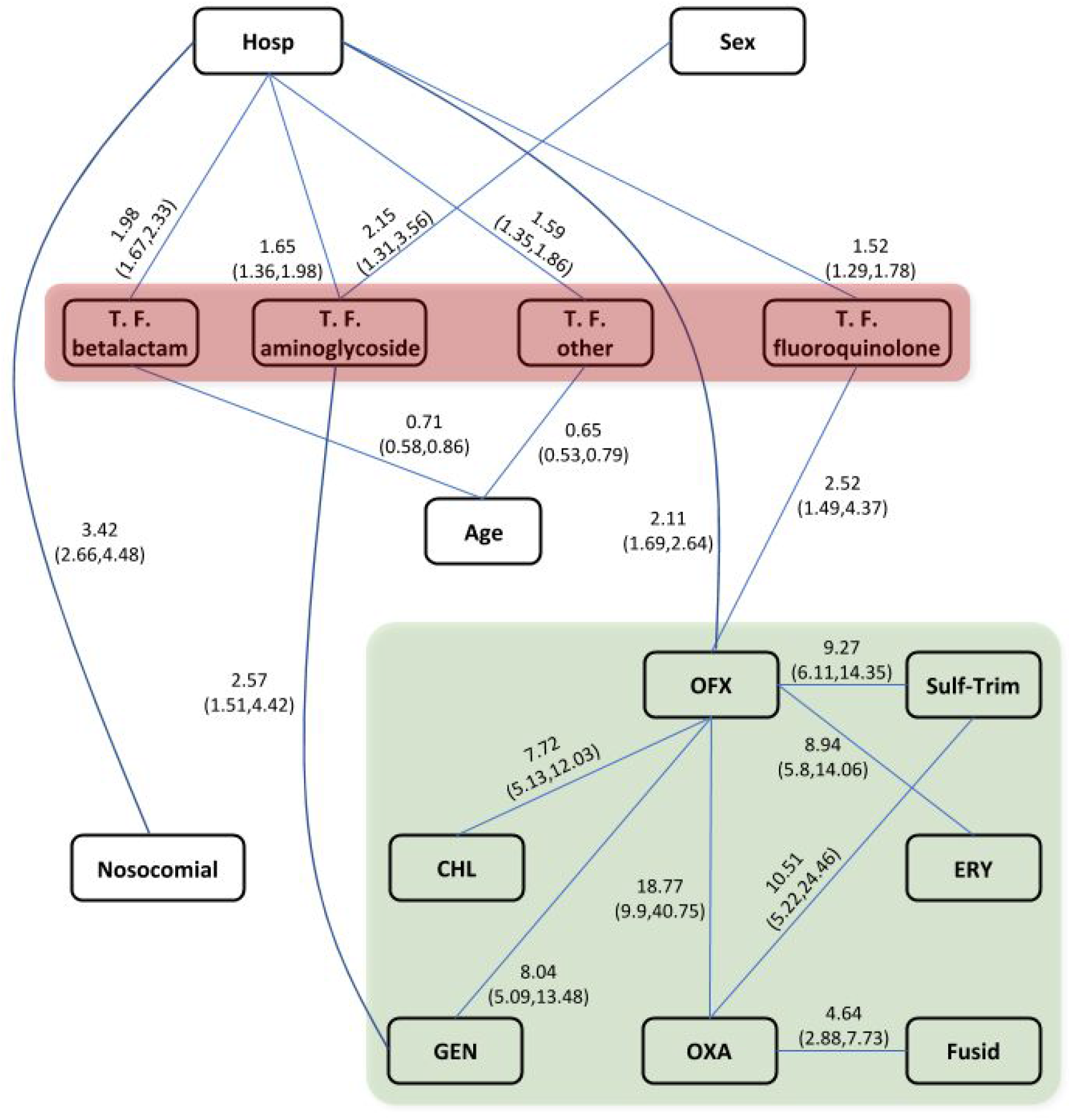
Directed acyclic graph of the final reduced model for CoNS. Arcs denote direct links between variables, and adjacent values are ORs derived from the model, along with their 95% credible intervals (see Table S6). Nodes with antibiotic names denote resistance to those antibiotics (enclosed in the light grey rectangle [green in the color online version]). T.F. prefix denotes variables indicating having taken an antibiotic from the given class in the prior year (enclosed in the dark grey rectangle [red in the color online version]); Hosp = log(days hospitalized + 1); Poly = polymicrobial culture; CHL = chloramphenicol; ERY = erythromycin; Fusid = fusidic acid; GEN = gentamicin; OFX = ofloxacin; OXA = oxacillin; Sulf-Trim = sulfamethoxazole-trimethoprim; Sex=male.

To summarize the analyses across the five bacterial species examined, we constructed a matrix of all possible arcs present in any of the five models (see Supplementary Figure 1). Within each cell, we present the number of models which contained the corresponding arc over the number of analyses which could have contained that arc (that is, counting datasets in which both variables were present). For ease of interpretation, variables which appeared in only a single model were then omitted (Figure 6). Importantly, the signs of all coefficients, when arcs were found, were consistent throughout the analysis of all five datasets. That is, no link between variables was found negative for a certain bacterial species and positive in another.

**Figure 6:**
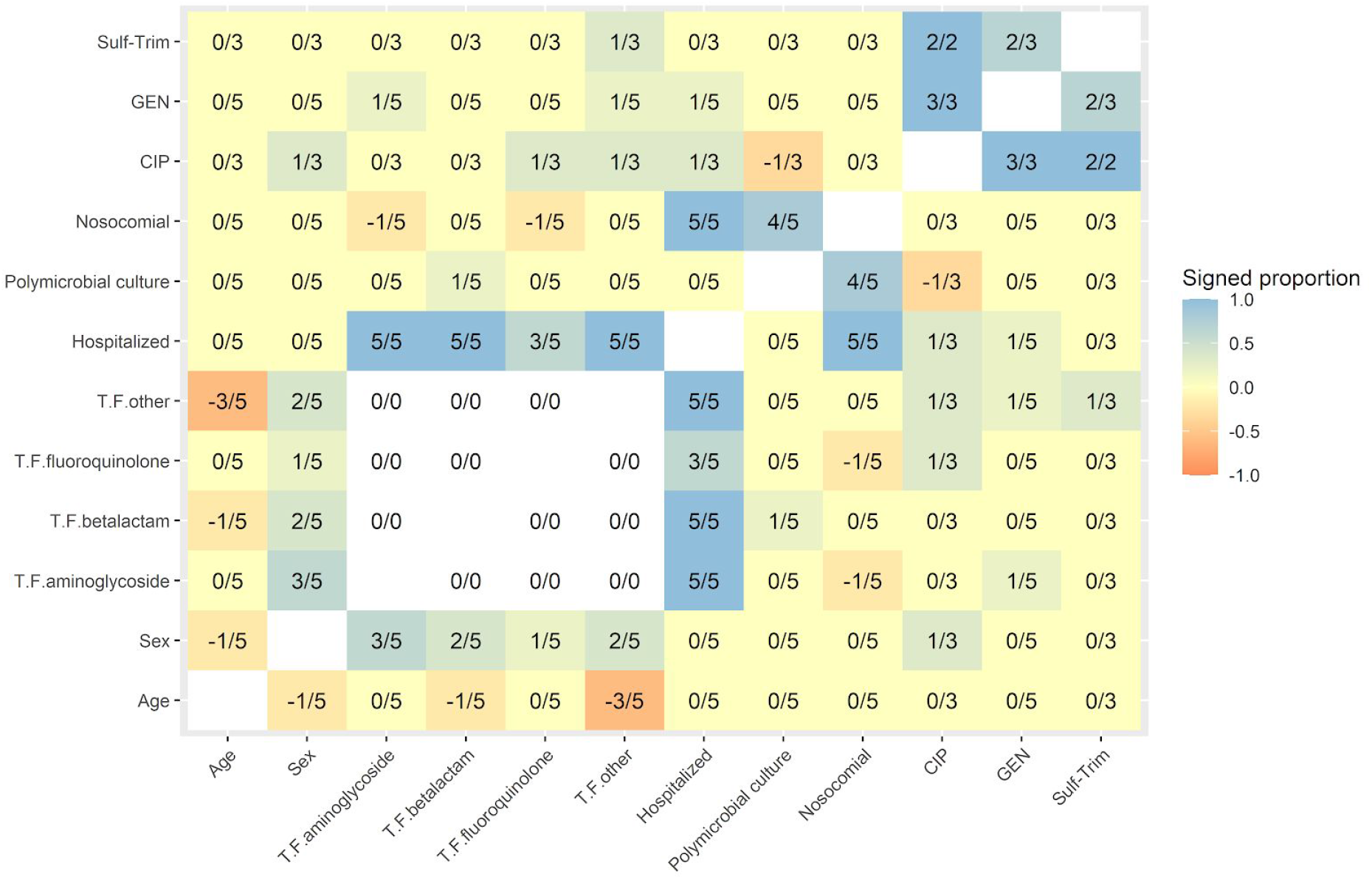
Consistency matrix showing proportion of models which contain a given arc. Each cell presents the proportion of models containing both variables and having a direct link between them (negative representing OR < 1). Shading represents the proportion of models containing the arc (in color in the online version). All signs of links were consistent between the datasets and shown in the figure. Variables which appear in only one of the 5 bacterial species datasets were omitted for clarity (but are found in Figure S1). Sex = male; T.F. prefix denotes having taken an antibiotic from the given class; Hosp = log(days hospitalized + 1); Poly = polymicrobial culture; CIP = ciprofloxacin; GEN = gentamicin; Sulf-Trim = sulfamethoxazole-trimethoprim.

The most important results from our analyses were the direct connections found between resistance to different antibiotics. **These were consistently positive throughout all antibiotics and all bacterial species (Figures 1-5)**. This implies that even when accounting for the possible connections between the different variables, acquisition of resistance to a certain antibiotic only served to enhance the probability of resistance to other antibiotics in our data.

Interestingly, we found several robust links between antibiotics of different classes. Direct links from different antibiotic classes to gentamicin were especially prevalent. Gentamicin and sulfamethoxazole-trimethoprim were directly linked in the *E. coli* (Figure 1) and *P. mirabilis* (Figure 4) datasets. Furthermore, gentamicin resistance was directly linked to resistance to several betalactam antibiotics. It was directly linked to both resistance to ceftazidime and imipenem (Figure 3) in the *P. aeruginosa* dataset and to cefuroxime (Figure 4) in the *P. mirabilis* dataset. Gentamicin resistance was also directly linked to resistance to ciprofloxacin, a fluoroquinolone, in the *E. coli, K. pneumoniae*, and *P. mirabilis* datasets (Figures 1, 2, and 4, respectively). Additionally, sulfamethoxazole-trimethoprim, was directly linked to ciprofloxacin in both datasets where they were tested *(E. coli*, Figure 1; *P. mirabilis*, Figure 4).

In all bacterial species we found a positive, direct link between having taken a betalactam, aminoglycoside, or "other” (i.e., any antibiotic not a member of the three classes which were most frequently used) antibiotic in the prior year, with days hospitalized in the prior year. In addition, three out of five of the analyses found a relationship between days hospitalized in the prior year and having taken a fluoroquinolone antibiotic. These relationships are somewhat expected, as more days spent at the hospital likely positively associated with having received antibiotic treatment, and vice versa.

Age and sex were only directly linked in the *P. mirabilis* dataset (Figure 4), with females being older at the time of the bacterial infection. When analyzing *E. coli* (Figure 1), *K. pneumoniae* (Figure 2), and CoNS (Figure 5), we found that older people were less likely to be prescribed an antibiotic in the "other” category. Older people were also less likely to be prescribed a betalactam in the CoNS model (Figure 5). Males were more likely to be prescribed aminoglycosides *(E. Coli*, Figure 1; *P. mirabilis*, Figure 4; CoNS, Figure 5), betalactams and other antibiotics *(E. Coli*, Figure 1; *P. mirabilis*, Figure 4), and fluoroquinolones *(E. Coli*, Figure 1). Additionally, males had infections with increased resistance to ciprofloxacin in the *E. coli* dataset (Figure 1), and to ceftazidime in the *P. aeruginosa* dataset (Figure 3). Resistance to cefuroxime was also positively linked to age in the *E. coli* dataset.

Having a nosocomial infection was directly linked to the presence of multiple bacteria in the culture in nearly all bacterial species, as has been shown,^31,32^ with CoNS being the one exception. This is plausible, as CoNS are often isolated as a result of culture contamination,^33^ and their presence might be associated with a decreased probability of the culture yielding other bacterial species. Correspondingly, the presence of additional bacterial species in a sample containing CoNS was less frequent than in all other bacterial species (7%, see Table 1). Nosocomial infection was also related to the number of days hospitalized during the prior year in all five bacterial species. This is expected, since acquiring a nosocomial infection by definition entails that the patient was hospitalized beforehand. Additionally, nosocomial infections were directly linked to having taken a fluoroquinolone in the *P. aeruginosa* dataset (Figure 3) and having taken an aminoglycoside in the *E. coli* dataset (Figure 1). Polymicrobial cultures were only directly linked to decreased resistance to one drug (ciprofloxacin) in only one bacterial species (*P. mirabilis*, Figure 4).

## Discussion

We have performed an additive Bayesian network analysis in an attempt to uncover the underlying dependency structure among patient covariates, measures of antibiotic use, and different antibiotic resistance types, in cultures of five bacterial species. Such an analysis has the advantage of not only shedding light on the relationships between all pairs of variables examined, but can identify which variables are directly linked rather than only having indirect associations.

All the connections between resistance to antibiotics identified by our statistical models, in all examined bacterial species, indicated positive associations. That is, resistance to a certain antibiotic was never found to decrease resistance to other antibiotics. Furthermore, some of the associations between resistance to different antibiotics were so strong they could not be included in the ABN models without causing numerical issues. This is expected with very similar antibiotics, but surprisingly appeared between antibiotics of different classes as well (Table 2).

Altogether, our results appear at odds with recent experimental findings. Although cross-resistance is prevalent in experimental settings, even between different classes of antibiotics, collateral sensitivity is also often observed.^10–13,34^ Nevertheless, as opposed to experimental data, clinical findings usually identify positive associations between resistance of different antibiotics.^35^ However, the reasons for the discrepancy between the experimental and clinical results may be hard to elucidate due to the potential presence of confounders in observational, clinical data. By using ABN modelling, we made a step towards resolving this problem and obtained estimates that reflect innate cross-resistance patterns rather than heavily confounded associations. Hence, our results strengthen the notion that at least some of the cross-resistance patterns observed in clinical settings relate to innate bacterial mechanisms of resistance acquisition. For instance, the direct cross-resistance we identified between fluoroquinolones and macrolides, between beta-lactams and aminoglycosides, and between aminoglycosides and quinolones, have been previously demonstrated in other clinical settings.^36–38^ Some other direct links in our models conform with prior biological and clinical knowledge: infections occurring within hospitals are expected to have a higher chance of containing several organisms than community acquired infections, and indeed nosocomial infections have been shown to be associated with polymicrobial cultures.^39^ Moreover, resistance of *E.coli* to ciprofloxacin and of various gram-negative bacteria to third generation cephalosporins has been shown to be higher in males.^40,41^ Although hypotheses regarding the reasons for this association have been suggested, e.g., different ages of hospitalizations due to urinary tract infections or avoidance of certain antibiotics during pregnancy, its definitive reason remains unclear.

Our results could have practical implications on strategies of antibiotic treatment. Knowledge of predictors for antibiotic resistance, and in particular for cross-resistance and previous antibiotic usage, is important for tailoring efficient antibiotic treatment regimes to patients. Combination therapy, for example, where a patient is treated with multiple antibiotics simultaneously, has been shown to yield beneficial results to mitigate resistance.^37,42,43^ However, the efficiency of combination therapy can be hampered by acquisition of multiple antibiotic resistance in the infecting bacteria. By having estimates of risks for cross-resistance based on patient demographics and previous antibiotic usage, we can mitigate this risk and seek to treat with combinations of antibiotics which have lower probability of cross-resistance. Alternatively, if such drug combinations are unavailable, attention could be focused on identifying new drugs meeting those needs in the future.

Moreover, our results could be used to suggest appropriate drugs for population-level alternating or cycling regimes of antibiotic therapy, which have been hypothesized in various settings as a way to reduce emergence of antibiotic resistance.^44–46^ Suggestions for selecting such drugs using estimates inferred from ABNs take into account the clinical characteristics of the patients and circulating bacterial strains. Hence, ABNs can identify patterns of cross-resistance which might be different than those obtained *in vitro* and can provide additional information about the potential efficiency of different antibiotic regimes. For example, alternating between gentamicin and cefuroxime has been offered as a treatment strategy against *E.coli* based on *in vitro* estimates,^10^ but our results suggest that these are directly linked and that ampicillin should have lower cross-resistance with gentamicin (Table 2 and Figure 1).

However, a limitation of our study is that our data lack previous exposure to antibiotics outside the hospital. Potentially, patients could have been exposed to antibiotic treatment in the community rather than during hospitalization, introducing noise into our estimates of the effects of previous antibiotic usage. However, our previous study on a similar dataset showed that prior antibiotic use in the hospital is a strong predictor for antibiotic resistance which can yield high accuracy predictions even without outpatient antibiotic use.^5^ Additionally, the consistency of our estimates between bacterial species, as well as relatively narrow coverage of the estimated CIs, indicate that our results should be robust to these missing data.

To conclude, our study provides estimates for cross-resistance between different antibiotics in bacterial pathogens isolated in clinical settings. By using an ABN model, we control for confounders and provide results for direct and indirect links between different patient characteristics and antibiotic resistance. Although our analysis does not provide direct causal effect estimates, it presents a step towards studying causal effects on antibiotic resistance patterns. Revealing the dependency structures of multiple variables simultaneously instead of analysing associations affecting a single dependent variable should provide more robust and less spurious relationships between variables. Hence, our results should be of utility in decision making regarding antibiotic treatments strategies.

## Data Availability

Data are proprietary.

## References

1. Friedman ND, Temkin E, Carmeli Y. The negative impact of antibiotic resistance. Clin Microbiol Infect 2016; 22: 416–22.

2. Rodríguez-Rojas A, Rodríguez-Beltrán J, Couce A, Blázquez J. Antibiotics and antibiotic resistance: a bitter fight against evolution. Int J Med Microbiol IJMM 2013; 303: 293–7.

3. Anon. Antibiotic Resistance Threats in the United States. Atlanta, GA, USA: Department of Health and Human Services, CDC; 2019. Available at: http://dx.doi.org/10.15620/cdc:82532.

4. Yelin I, Snitser O, Novich G, et al. Personal clinical history predicts antibiotic resistance of urinary tract infections. Nat Med 2019; 25: 1143–52.

5. Lewin-Epstein O, Baruch S, Hadany L, Stein G, Obolski U. Predicting antibiotic resistance in hospitalized patients by applying machine learning to electronic medical records. medRxiv 2020: 2020.06.03.20120535.

6. Chatterjee A, Modarai M, Naylor NR, et al. Quantifying drivers of antibiotic resistance in humans: a systematic review. Lancet Infect Dis 2018; 18: e368–78.

7. Anon. CROSS RESISTANCE TO ANTIBIOTICS. J Am Med Assoc 1952; 148: 470–1.

8. Obolski U, Dellus-Gur E, Stein GY, Hadany L. Antibiotic cross-resistance in the lab and resistance co-occurrence in the clinic: Discrepancies and implications in E.coli. Infect Genet Evol J Mol Epidemiol Evol Genet Infect Dis 2016; 40: 155–61.

9. Blair JMA, Webber MA, Baylay AJ, Ogbolu DO, Piddock LJV. Molecular mechanisms of antibiotic resistance. Nat Rev Microbiol 2015; 13: 42–51.

10. Imamovic L, Sommer MOA. Use of Collateral Sensitivity Networks to Design Drug Cycling Protocols That Avoid Resistance Development. Sci Transl Med 2013; 5: 204ra132–204ra132.

11. Pál C, Papp B, Lázár V. Collateral sensitivity of antibiotic-resistant microbes. Trends Microbiol 2015; 23: 401–7.

12. Lázár V, Nagy I, Spohn R, et al. Genome-wide analysis captures the determinants of the antibiotic cross-resistance interaction network. Nat Commun 2014; 5: 4352.

13. Munck C, Gumpert HK, Wallin AIN, Wang HH, Sommer MOA. Prediction of resistance development against drug combinations by collateral responses to component drugs. Sci Transl Med 2014; 6: 262ra156–262ra156.

14. Kratzer G, Lewis FI, Comin A, Pittavino M, Furrer R. Additive Bayesian Network Modelling with the R Package abn. ArXiv Prepr ArXiv191109006 2019.

15. Lewis FI, Ward MP. Improving epidemiologic data analyses through multivariate regression modelling. Emerg Themes Epidemiol 2013; 10: 4.

16. Kratzer G, Pittavino M, Lewis FI, Furrer R. abn: an R package for modelling multivariate data using additive Bayesian networks. 2019. Available at: https://CRAN.R-project.org/package=abn.

17. Ruchti S, Meier AR, Würbel H, Kratzer G, Gebhardt-Henrich SG, Hartnack S. Pododermatitis in group housed rabbit does in Switzerland—Prevalence, severity and risk factors. Prev Vet Med 2018; 158: 114–21.

18. Ludwig A, Berthiaume P, Boerlin P, Gow S, Léger D, Lewis FI. Identifying associations in Escherichia coli antimicrobial resistance patterns using additive Bayesian networks. Prev Vet Med 2013; 110: 64–75.

19. Hartnack S, Odoch T, Kratzer G, et al. Additive Bayesian networks for antimicrobial resistance and potential risk factors in non-typhoidal Salmonella isolates from layer hens in Uganda. BMC Vet Res 2019; 15: 212.

20. Hidano A, Yamamoto T, Hayama Y, et al. Unraveling Antimicrobial Resistance Genes and Phenotype Patterns among Enterococcus faecalis Isolated from Retail Chicken Products in Japan. PLoS ONE 2015; 10. Available at: https://www.ncbi.nlm.nih.gov/pmc/articles/PMC4363150/. Accessed February 17, 2020.

21. Waterhouse M, Morton A, Mengersen K, Cook D, Playford G. Role of overcrowding in meticillin-resistant Staphylococcus aureus transmission: Bayesian network analysis for a single public hospital. J Hosp Infect 2011; 78: 92–6.

22. R Core Team. R: A Language and Environment for Statistical Computing. Vienna, Austria: R Foundation for Statistical Computing; 2019. Available at: https://www.R-project.org/.

23. Friedman N, Goldszmidt M, Wyner A. Data Analysis with Bayesian Networks: A Bootstrap Approach. *ArXiv13016695 Cs Stat* 2013. Available at: http://arxiv.org/abs/1301.6695. Accessed April 21, 2020.

24. Plummer M. JAGS: A program for analysis of Bayesian graphical models using Gibbs sampling. Proc 3rd Int Workshop Distrib Stat Comput 2003; 124: 1–10.

25. Lewis FI, McCormick BJJ. Revealing the Complexity of Health Determinants in Resource-poor Settings. Am J Epidemiol 2012; 176: 1051–9.

26. Poon AFY, Lewis FI, Pond SLK, Frost SDW. Evolutionary Interactions between N-Linked Glycosylation Sites in the HIV-1 Envelope. PLoS Comput Biol 2007; 3. Available at: https://www.ncbi.nlm.nih.gov/pmc/articles/PMC1779302/. Accessed February 17, 2020.

27. Poon AFY, Lewis FI, Pond SLK, Frost SDW. An Evolutionary-Network Model Reveals Stratified Interactions in the V3 Loop of the HIV-1 Envelope. PLoS Comput Biol 2007; 3. Available at: https://www.ncbi.nlm.nih.gov/pmc/articles/PMC2082504/. Accessed February 17, 2020.

28. Poon AFY, Lewis FI, Frost SDW, Kosakovsky Pond SL. Spidermonkey: rapid detection of co-evolving sites using Bayesian graphical models. Bioinformatics 2008; 24: 1949–50.

29. Lycett SJ, Ward MJ, Lewis FI, Poon AFY, Kosakovsky Pond SL, Brown AJL. Detection of Mammalian Virulence Determinants in Highly Pathogenic Avian Influenza H5N1 Viruses: Multivariate Analysis of Published Data. J Virol 2009; 83: 9901–10.

30. Milns I, Beale CM, Smith VA. Revealing ecological networks using Bayesian network inference algorithms. Ecology 2010; 91: 1892–9.

31. Siegman-Igra Y, Kulka T, Schwartz D, Konforti N. Polymicrobial and monomicrobial bacteraemic urinary tract infection. J Hosp Infect 1994; 28: 49–56.

32. Weinstein MP, Reller LB, Murphy JR. Clinical importance of polymicrobial bacteremia. Diagn Microbiol Infect Dis 1986; 5: 185–96.

33. Altindis M, Koroglu M, Demiray T, et al. A Multicenter Evaluation of Blood Culture Practices, Contamination Rates, and the Distribution of Causative Bacteria. Jundishapur J Microbiol 2016; 9. Available at: https://www.ncbi.nlm.nih.gov/pmc/articles/PMC4834024/. Accessed April 22, 2020.

34. Adamus-Białek W, Wawszczak M, Arabski M, et al. Ciprofloxacin, amoxicillin, and aminoglycosides stimulate genetic and phenotypic changes in uropathogenic Escherichia coli strains. Virulence 2019; 10: 260–76.

35. Chang H-H, Cohen T, Grad YH, Hanage WP, O’Brien TF, Lipsitch M. Origin and Proliferation of Multiple-Drug Resistance in Bacterial Pathogens. Microbiol Mol Biol Rev 2015; 79: 101–16.

36. Hwang TJ, Hooper DC. Association between fluoroquinolone resistance and resistance to other antimicrobial agents among Escherichia coli urinary isolates in the outpatient setting: a national cross-sectional study. J Antimicrob Chemother 2014; 69: 1720–2.

37. Monedero I, Caminero JA. Management of multidrug-resistant tuberculosis: an update. Ther Adv Respir Dis 2010; 4: 117–27.

38. Tsukamoto N, Ohkoshi Y, Okubo T, et al. High Prevalence of Cross-Resistance to Aminoglycosides in Fluoroquinolone-Resistant Escherichia coli Clinical Isolates. Chemotherapy 2013; 59: 379–84.

39. Yo C-H, Hsein Y-C, Wu Y-L, et al. Clinical predictors and outcome impact of community-onset polymicrobial bloodstream infection. Int J Antimicrob Agents 2019; 54: 716–22.

40. Lee DS, Choe H-S, Kim HY, et al. Role of age and sex in determining antibiotic resistance in febrile urinary tract infections. Int J Infect Dis 2016; 51: 89–96.

41. Livermore DM, Nichols T, Lamagni TL, Potz N, Reynolds R, Duckworth G. Ciprofloxacin-resistant Escherichia coli from bacteraemias in England; increasingly prevalent and mostly from men. J Antimicrob Chemother 2003; 52: 1040–2.

42. REX Consortium. Heterogeneity of selection and the evolution of resistance. Trends Ecol Evol 2013; 28: 110–8.

43. Vandamme A-M, Camacho RJ, Ceccherini-Silberstein F, et al. European Recommendations for the Clinical Use of HIV Drug Resistance Testing: 2011 Update. AIDS Rev 2011; 13. Available at: https://pubmed.ncbi.nlm.nih.gov/21587341/. Accessed May 23, 2020.

44. Kim S, Lieberman TD, Kishony R. Alternating antibiotic treatments constrain evolutionary paths to multidrug resistance. Proc Natl Acad Sci U S A 2014; 111: 14494–9.

45. Obolski U, Hadany L. Implications of stress-induced genetic variation for minimizing multidrug resistance in bacteria. BMC Med 2012; 10: 89.

46. Obolski U, Stein GY, Hadany L. Antibiotic Restriction Might Facilitate the Emergence of Multi-drug Resistance. PLOS Comput Biol 2015; 11: e1004340.

